# Previous psychopathology predicted severe COVID-19 concern, anxiety and PTSD symptoms in pregnant women during lockdown in Italy

**DOI:** 10.1101/2020.08.26.20182436

**Authors:** Claudia Ravaldi, Valdo Ricca, Alyce Wilson, Caroline Homer, Alfredo Vannacci

## Abstract

Italy was the first COVID-19 pandemic epicenter among European countries and established a period of full lockdown, consisting of travel bans, mandatory staying at home and temporary closure of non-essential businesses. Similar measures are known risk factors for psychological disturbances in the general population, still very little is known about their impact on pregnant women’s mental health during COVID-19 pandemic.

The national survey “COVID-19 related Anxiety and StreSs in prEgnancy, poSt-partum and breaStfeeding” (COVID-ASSESS) was conducted during the first month of full lockdown in Italy. The questionnaire was specifically developed to examine COVID-19 concerns and included the psychometric tests NSESSS for PTSD and STAI-Y for anxiety. A multivariable logistic regression model was fitted to explore the association of the concern, anxiety and PTSD symptoms with age, gestational weeks, parity, days of lockdown, assisted reproductive technology use, psychopathological history and previous perinatal losses.

Out of 1015 pregnant women reached, 737 (72.6%) fully answered the questionnaire; no woman reported a COVID-19 infection. Median age was 34.4 years [quartiles 31.7, 37.2], median days in lockdown were 13.1 [11.0, 17.0], median gestational weeks were 27.8 [19.8, 34.0]. Clinically significant PTSD symptoms were present in 75 women (10.2%, NSESSS cut-off 24) and clinically significant anxiety symptoms were present in 160 women (21.7%, STAI-Y1 cut-off 50). Women were less worried about their own health than the health of their baby and of their elderly relatives. Previous anxiety predicted higher concern and PTSD symptoms; previous depression and anxiety were independently associated with current PTSD symptoms.

## Background

The newly identified Coronavirus SARS-CoV-2, responsible for the associated respiratory infection designated COVID-19, was first identified in December 2019 in Wuhan, China, and spread to Europe and worldwide within months. Italy was the first pandemic epicenter among European countries. On 9 March 2020, the Government of Italy established a period of full lockdown, consisting of travel bans, mandatory staying at home for all (except for emergencies, health problems or regulated shopping for bare necessities) and temporary closure of non-essential shops and businesses, that lasted until 3 May 2020. These measures, initially considered strict and controversial, eventually controlled SARS-CoV-2 transmission in all Italian regions. In the following months, similar measures have been implemented in many European [1] and other countries globally to control the pandemic. Quarantine and self-isolation are now common practices in many countries to prevent transmission of the virus. In non-Covid times quarantine and social isolation are a well-known risk factors for psychological and psychiatric disturbances in the general population [2], particularly for children and adolescents, the elderly, those from lower socio-economic groups, females and people with pre-existing mental health conditions [3]. However, very little is known about the impact on the mental health of pregnant women during the COVID-19 pandemic.

## Methods

During the first weeks of full lockdown in Italy (18-31 March 2020), we conducted a national survey “COVID-19 related Anxiety and StreSs in prEgnancy, poSt-partum and breaStfeeding” (COVID-ASSESS) to investigate the psychological impact of the pandemic and containment measures on pregnant women. The research methods are described elsewhere [4], Briefly, a cross-sectional, web-based study was conducted using an online questionnaire. Participants were recruited using snowball sampling via social networks with a snowball technique and sponsored social network advertisements. Approval for the study was obtained from the ethics committee of Florence University. Participants self-selected to complete the survey, participation was voluntary and all participants gave their consent in an online form. The online tool - known as the COVID-ASSESS questionnaire - comprised of a sociodemographic section, a specifically developed survey to examine concerns related to the COVID-19 pandemic and two validated psychometric tests: NSESSS for PTSD [5] and STAI-Y [6].

Categorical variables were reported as n (%), continuous variables as median [quartiles; range min-max]; A multivariable logistic regression model was fitted to explore the association of the selected outcomes (concern, anxiety and Post Traumatic Stress Disorder (PTSD) symptoms) with age, gestational weeks, parity, days of lockdown, assisted reproductive technology use, psychopathological history and previous perinatal losses. Analyses were conducted with Stata/IC 16.1 (StataCorp), a value of p<0.01 was considered statistically significant. Graphs were plotted with Tableau Desktop 2020.1 (Tableau Software, LLC).

## Results

Although response rates could not be exactly quantifiable due to the self-selected and non-probabilistic nature of the sample, 737 out of 1015 pregnant women who logged in to the web-based survey, fully answered the COVID-ASSESS questionnaire (72.6%); no woman reporting having experienced a COVID-19 infection. The median age was 34.4 years [31.7, 37.2; range 18.4 - 47.4], median days in lockdown at the time of interview were 13.1 [11.0, 17.0; range 7.8 - 27.5]; 263 women (35.7%) were at their first pregnancy (no previous childbirth, no previous pregnancy loss), median gestational weeks were 27.8 [19.8, 34.0; range 4.7 - 42.5] and the distribution of pregnancy trimesters was: first trimester 67 (9.1%), second trimester 309 (41.9%), third trimester 361 (49.0%). Assisted reproductive technology was reported by 45 (5.8%) women and 277 women (37.6%) reported at least one previous pregnancy or postnatal loss [miscarriages 204 (27.7%); termination of pregnancy 75 (10.2%); stillbirths or neonatal/infant losses 36 (4.8%)].

Previous psychopathological diagnoses were reported by 293 women (39.8%), in particular: anxiety 241 (32.7%), depression 69 (9.3%), bipolar disorder 4 (0.5%), Obsessive Compulsive Disorder (OCD) 9 (1.2%), and eating disorders 46 (6.2%). Women with a history of anxiety were significantly more concerned about COVID-19. On a Likert scale from 0 to 3, women with no previous psychopathological diagnosis scored a median of 2.28 [2.00; 2.71], vs 2.43 [2.14; 2.71] of women with a psychopathological history (p=0.003); the difference was more pronounced regarding health-related issues (figure 1A, blue bars), than society-related issues (figure 1A, grey bars). Consistently, a previous diagnosis of anxiety was the only factor able to significantly predict high levels of concern in the multivariate analysis (OR 1.85; CI 1.16, 2.95).

**Figure 1.**
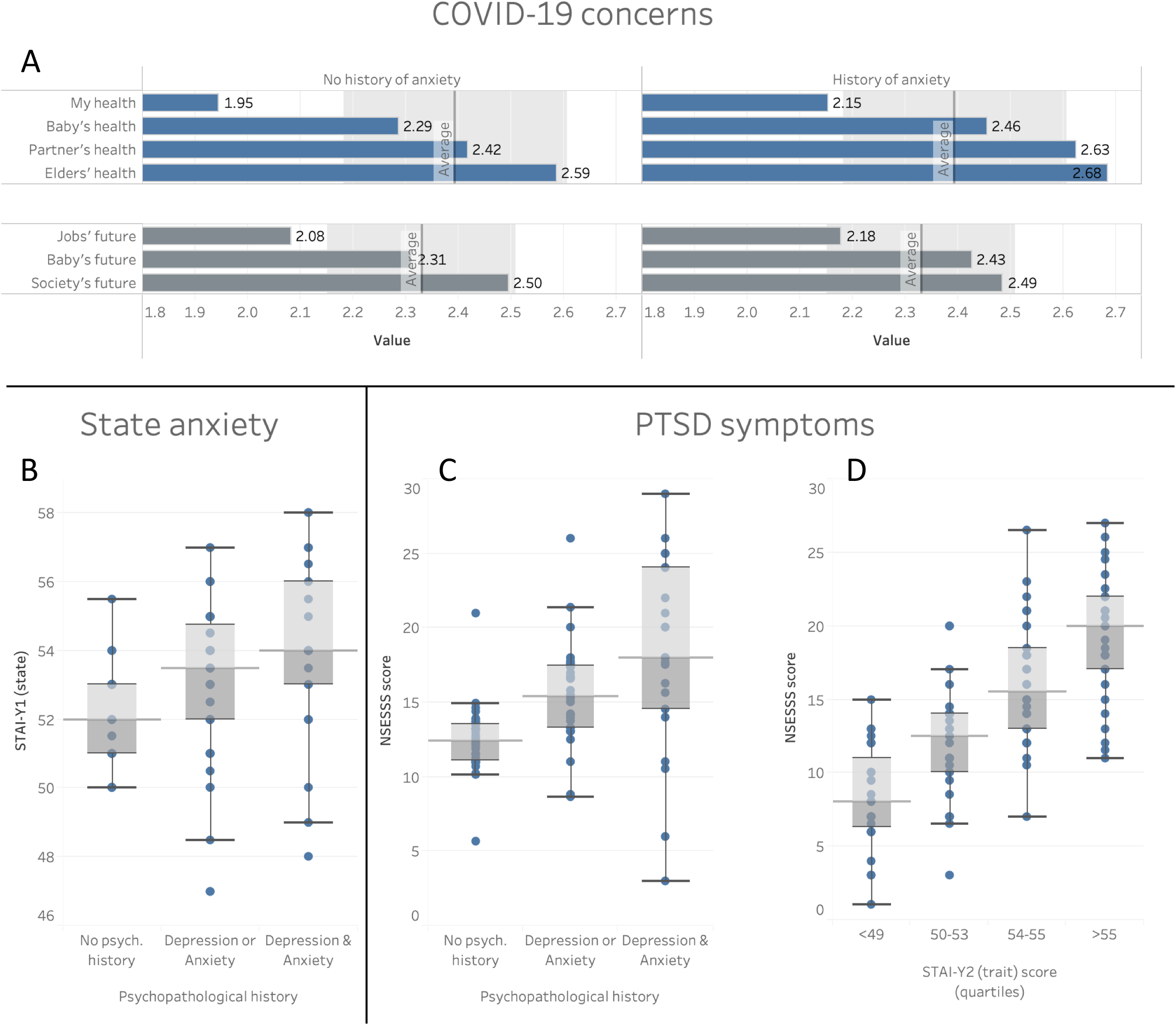
Level of concern of women according to history of psychological distress (panel A); state anxiety according to psychopathological history (panel B); post traumatic stress disorder (PTSD) symptoms according to psychopathological history (panel C) and anxiety trait (panel D).

With regard to psychopathological outcomes, clinically significant PTSD symptoms were present in 75 women (10.2%) using a NSESSS cut-off of 24 [7] and clinically significant anxiety symptoms were present in 160 women (21.7%) using a STAI-Y1 cut-off of 50 [8]. The most important factors correlated to high levels of psychopathology were previous diagnoses of anxiety and/or depression. Women with a history of anxiety or depression showed significantly higher levels of anxiety, measured as STAI-Y1 score (figure 1B; no previous psychopathology 52 [49; 55] vs previous depression or anxiety 53 [51; 56] vs previous depression and anxiety 54 [51; 56]; p=0.0001).

Finally, women with a history of anxiety or depression showed significantly more PTSD symptoms (figure 1C; no previous psychopathology 12 [7; 17], previous depression or anxiety 16 [10; 22], previous depression and anxiety 19 [11; 25]; p=0.0001). Previous depression and anxiety diagnoses were also independently associated with the risk of developing PTSD symptoms in the multivariate analysis (p=0.004): previous depression OR 2.31 (1.18, 4.50), previous anxiety OR 2.30 (1.38, 3.82), previous depression and anxiety OR 5.66 (2.56, 12.54). Consistently, women with an anxiety trait (measured by STAI-Y2) showed higher levels of PTSD symptoms (figure 1D; Rsq=0.11, p=0.0001).

Results of this nation-wide survey conducted during the first weeks of lockdown in Italy show that SARS-CoV-2 negative pregnant women were very concerned about COVID-19 and showed a high prevalence of anxiety and post-traumatic stress disorder symptoms. Higher levels of concern and psychological distress were significantly associated with a previous psychopathological diagnosis. In particular, women with self-reported history of anxiety and/or depression were significantly more concerned about COVID-19 and were at a higher risk of developing symptoms of anxiety and post-traumatic stress disorder.

These findings have immediate relevance for healthcare professionals caring for pregnant and postnatal women during the COVID-19 pandemic. When taking a medical history, it is critical that care givers ask women about any history of anxiety and/or depression as this seems to be the most important factor in predicting COVID-19 associated distress and psychopathology during pregnancy. Healthcare professionals caring for pregnant and postnatal women during current or future public health crises should be aware that the simple act of asking women about previous psychopathological diagnoses can be a simple and useful way to identify patients in need of particular care and attention, in order to reduce the risk of adverse psychological outcomes, such as PTSD and postpartum depression.

## Data Availability

Raw data is available in Mendeley repository

